# Paradoxical Effects of Depression on Psoriatic Arthritis Outcomes in a Combined Psoriasis-Psoriatic Arthritis Center

**DOI:** 10.1101/2022.09.19.22280104

**Authors:** Rebecca H. Haberman, Seungha Um, Sydney Catron, Alan Chen, Eileen Lydon, Malavika Attur, Andrea L. Neimann, Soumya Reddy, Andrea Troxel, Samrachana Adhikari, Jose U. Scher

**Author notes:** Corresponding author: Rebecca Haberman. **Disclosures:** RHH has served as a consultant for Janssen. ALN declares that she has served as a consultant for Janssen, UCB, AbbVie, BMS and her immediate family member owns shares of stock in J&J, Eli Lilly, AbbVie, and Pfizer. SR has served as a consultant for AbbVie/Abbott, Amgen, Novartis, Janssen, Pfizer. SA has received grants from Johnson and Johnson. JUS has served as a consultant for Janssen, Novartis, Pfizer, Sanofi, Amgen, UCB and AbbVie; and has received funding for investigator-initiated studies from Janssen and Pfizer. **Funding:** This study was funded by NIH/NIAMS (R01AR074500 to Scher, T32-AR-069515 to Haberman, Scher), Rheumatology Research Foundation, the National Psoriasis Foundation, The Beatrice Snyder Foundation, The Riley Family Foundation.

## Abstract

Psoriatic arthritis (PsA) is a chronic, complex, inflammatory arthritis that, when left untreated, can lead to erosions, deformities and decrease in quality of life. PsA is known to be associated with multiple comorbidities, including cardiovascular, metabolic and mental health syndromes, all of which can increase its overall morbidity and mortality. Here we describe a cohort of patients with PsA from an urban, tertiary care, and followed at a combined rheumatology-dermatology center. Depression was reported in 22.8% of the population, anxiety in 18%, and attention deficit hyperactivity disorder in 4%. Depression was more common in female participants (p<.001). At baseline, individuals with PsA and concomitant depression had similar tender and swollen joint counts and RAPID3 compared to those without depression, and had lower body surface area affected by psoriasis (p=.04). At year one, all patients had improvement in clinical outcomes. However, patients with depression had a significantly higher tender joint count compared to those without depression (p=.001), despite similar swollen joint count and body surface area. These observations underscore the importance of addressing depression and psychological distress as part of PsA treatment outcomes and points towards the need to address residual pain through co-adjuvant approaches.

## Introduction

Psoriatic arthritis (PsA) is a common immune mediated inflammatory arthritis with a prevalence on the rise in both the United States^1^ and across the world^2,3^. PsA has a complex and diverse phenotype, and affects up to 30% of patients with psoriasis. It involves multiple domains, including skin, nails, peripheral arthritis, axial disease, dactylitis, and enthesitis. The spectrum of inflammatory changes has a significant interindividual variability and can become evident either as isolated clinical manifestations or as part of a multi-domain syndrome. This phenotypic diversity also contributes to the significant underdiagnosis and undertreatment of PsA^4,5^, which are known to negatively impact disease outcomes.^6,7^.

Beyond its deleterious effects in skin and joints of patients ^8^, PsA is associated with multiple comorbid conditions (i.e., cardiovascular and metabolic disease), which contribute to increased rates of morbidity and mortality^9,10^. In addition to its physical toll, PsA can lead to decreased quality of life, high levels of psychosocial stress, and increased rates of unemployment and reduced productivity^11-13^.

Depression and anxiety also have a high prevalence amongst patients with psoriatic disease^14,15^. Previous studies have suggested that depression may impact the ability of patients to achieve remission in both rheumatoid arthritis and PsA^16^. However, the underlying effects of depression in psoriatic disease is neither well characterized nor commonly acknowledged in clinical practice. In fact, and despite significant progress in therapeutics over the last two decades, treatment of inflammation in PsA remains substandard and residual symptoms (most notably pain and fatigue) are not resolved in a significant proportion of patients, hampering their ability to achieve a state of remission or even low-disease activity^17^. We therefore hypothesized that the presence of depression significantly affects PsA outcomes and may be at least partially responsible for the endurance of these residual symptoms.

Here, we describe the prevalence and extent of psychiatric comorbidities in a longitudinal PsA cohort from an urban, tertiary care, combined clinical setting and further assessed the effects of depression on PsA outcomes.

## Methods

### Patients

Consecutive adult patients meeting CASPAR criteria for PsA^18^ were prospectively enrolled from the established New York University (NYU) Psoriatic Arthritis Center (PAC) from January 7, 2015 (inception of standardized electronic health record template) to December 3, 2020 into an observational, prospective cohort registry. During this time period, 527 patients were evaluated. Demographic and outcome information was extracted from clinical visits utilizing a PsA-specific template in the electronic health record (EHR; Epic^19^) and subsequently entered into a REDCap^20^ database. Baseline visit was defined as the first clinical interaction regardless of timepoint in disease duration or treatment. Patients were followed for up to 2 years from their baseline visit, for an average of 14.2 months [range 0.23-23.97 months]. All participating physicians are trained rheumatologists with specialization in psoriatic disease and practicing in a combined, dermatology-rheumatology clinic setting.

Demographic data, comorbidities, and family history were recorded. Mental health conditions (i.e., depression, anxiety, and attention deficit disorder (ADHD)) were defined by established diagnosis (patient report and/or ICD code) and/or use of psychiatric medication. Physical exam findings such as tender (TJC) and swollen joint counts (SJC), and body surface area affected by psoriasis (BSA) were recorded from clinical notes as assessed by their primary NYU PAC rheumatologist. Patient reported outcomes such as the Multidimensional Health Assessment Questionnaire (MDHAQ)^21^ and the resultant RAPID3^22^ were entered by the patients on iPads at each clinical visit as part of routine care and directly transmitted to the EHR.

### Statistical Analysis

Baseline characteristics of study participants during the first visit were summarized using frequency and proportion for categorical variables and mean and standard deviation for continuous variables. Statistical comparisons between the depressed and non-depressed groups, the primary exposure of interest, were performed using ANOVA and Wilcoxon rank sum tests for continuous variables, and chi-square tests for categorical variables. The association of depression with longitudinal PsA outcomes were assessed using mixed-effects Poisson regression models for tender and swollen joint counts and mixed-effects linear regression models for RAPID3 and BSA, adjusting for age, sex, race, medication use, comorbidities, and time since baseline. The model also included random intercepts for the individual patients, and interaction between depression and time. Estimates of rate ratios (RR) for the count outcomes and differences for the continuous outcomes along with 95% confidence intervals (CI), comparing the two exposure groups, were reported at the baseline, 1- and 2-year follow-up. All analyses were performed using R v4.1.2 software (R Foundation for Statistical Computing)^23^.

## Results

At the time of analysis, the NYU PAC cohort consisted of 527 patients with diagnosed PsA as defined by meeting CASPAR criteria. Patients were 46.7% female with a mean age of 49 years. The population was mostly white (79.7%), but did have a significant number of patients who identified as other races or ethnicities (**Table 1**). PsA phenotype was heterogeneous in clinical presentation, with 93.2% of patients showing peripheral joint involvement at any timepoint in disease course, 31.9% with axial involvement, 99.2% with skin involvement, 62.2% nail involvement, 35.1% entheseal involvement, and 33.6% dactylitis. Type of psoriasis was mostly in plaque form (75.1%) with 55.2% having nail involvement. Other types of psoriasis included: inverse (18.2%), guttate (4.7%), palmoplantar (3.6%), pustular (4.6%), and erythrodermic (0.4%).

**Table 1.** Baseline Characteristics.

| Characteristic | All<br>(n=527) | Depression<br>(n=120) | No depression<br>(n=407) | p-value<sup>#</sup> |
| --- | --- | --- | --- | --- |
| Age- mean (SD) | 49.0 (14) | 52.1 (15.2) | 48.1 (13.6) | 0.01 |
| Female- n (%) | 244 (46.7) | 74 (62.7) | 170 (42.0) | <0.001 |
| Race- n (%) |  |  |  | 0.86 |
| White | 420 (79.7) | 99 (82.5) | 321 (78.9) |  |
| Black | 7 (1.3) | 0 (0.0) | 7 (1.7) |  |
| Asian | 42 (8.0) | 4 (3.3) | 38 (9.3) |  |
| Other | 7 (1.3) | 2 (1.7) | 5 (1.2) |  |
| Hispanic ethnicity- n (%) | 51 (9.7) | 15 (12.5) | 36 (8.8) | 0.23 |
| Body Mass Index- mean<br>(SD) | 27.9 (5.9) | 28.5 (6.2) | 27.8 (5.7) | 0.24 |
| Comorbidities- n (%) |  |  |  |  |
| Anxiety | 95 (18.0) | 61 (50.8) | 34 (8.4) | <0.001 |
| ADHD | 21 (4.0) | 7 (5.8) | 14 (2.4) | 0.36 |
| Cardiovascular disease* | 171 (32.4) | 50 (41.7) | 121 (29.7) | 0.02 |
| Metabolic disease<sup>x</sup> | 68 (12.9) | 12 (15.8) | 49 (12.0) | 0.35 |
| Medication use- n (%) |  |  |  |  |
| Any | 386 (73.2) | 93 (77.5) | 293 (72.0) | 0.28 |
| Biologic | 223 (45.1) | 55 (47.8) | 168 (44.2) | 0.56 |

\*Cardiovascular disease includes hypertension, myocardial infarctions, hyperlipidemia, heart failure, angina, coronary artery disease, and stroke.
<sup>x</sup> Metabolic disease includes diabetes and obesity.

At the baseline visit, individuals had an average of 1.8 swollen joints (SD 3.1) and 3.1 tender joints (SD 4.6), with 31% of participants showing at least one area of active enthesitis. The average percent BSA was 2.6% (SD 5.3) with a mean RAPID3 score of 10.9 (SD 6.7). At baseline, 217 patients were previously seen at the PAC, 248 were new to the PAC but had previously been seen by other rheumatologists, and 62 were new to PAC and had never been seen by rheumatology. At the baseline visit, 73.2% of patients were already on a biologic or non-biologic DMARD, with 45.1% of them on anti-cytokine therapy or janus kinase inhibitors (JAKis) (**Table 1**). Patients had relatively high rates of comorbidities including hypertension, hyperlipidemia, diabetes, obesity, inflammatory bowel disease (IBD) and uveitis. Our cohort also displayed high rates of psychiatric disease including depression (22.8%), anxiety (18%), and ADHD (4%), with depression and anxiety co-existing in 11.6% of participants (**Table 1**). Of those patients with depression, 47.5% were on at least one medication classified as an antidepressant.

We then examined correlations between depression and disease measures at baseline. Participants with depression were more likely to be female (62.7% vs. 42.0%, P < 0.01) and older (52.1 years vs. 48.1 years, P=0.03) (**Table 1**). Compared to those without depression, patients with depression were more likely to have concomitant anxiety (50.8% vs. 8.4%, P < 0.01). They were also significantly more likely to have cardiovascular disease (including hypertension, hyperlipidemia, coronary artery disease, angina, and history of myocardial infraction and stroke; 41.7% vs. 29.7%, P=0.02) despite having comparable BMI measurements (28.5 vs. 27.8, P=0.24).

At baseline, individuals with depression had similar observed tender and swollen joint counts and RAPID3 scores, and a lower BSA (p=0.04) compared to those without depression (Figure 1). However, at one year, while both groups had improvement in their tender and swollen joint counts, patients with depression had a significantly higher TJC (p=0.004) despite similar SJC, RAPID3, and BSA compared to non-depressed individuals. By year two, although not achieving significance, those with depression had numerically higher TJC yet lower SJC.

**Figure 1.**
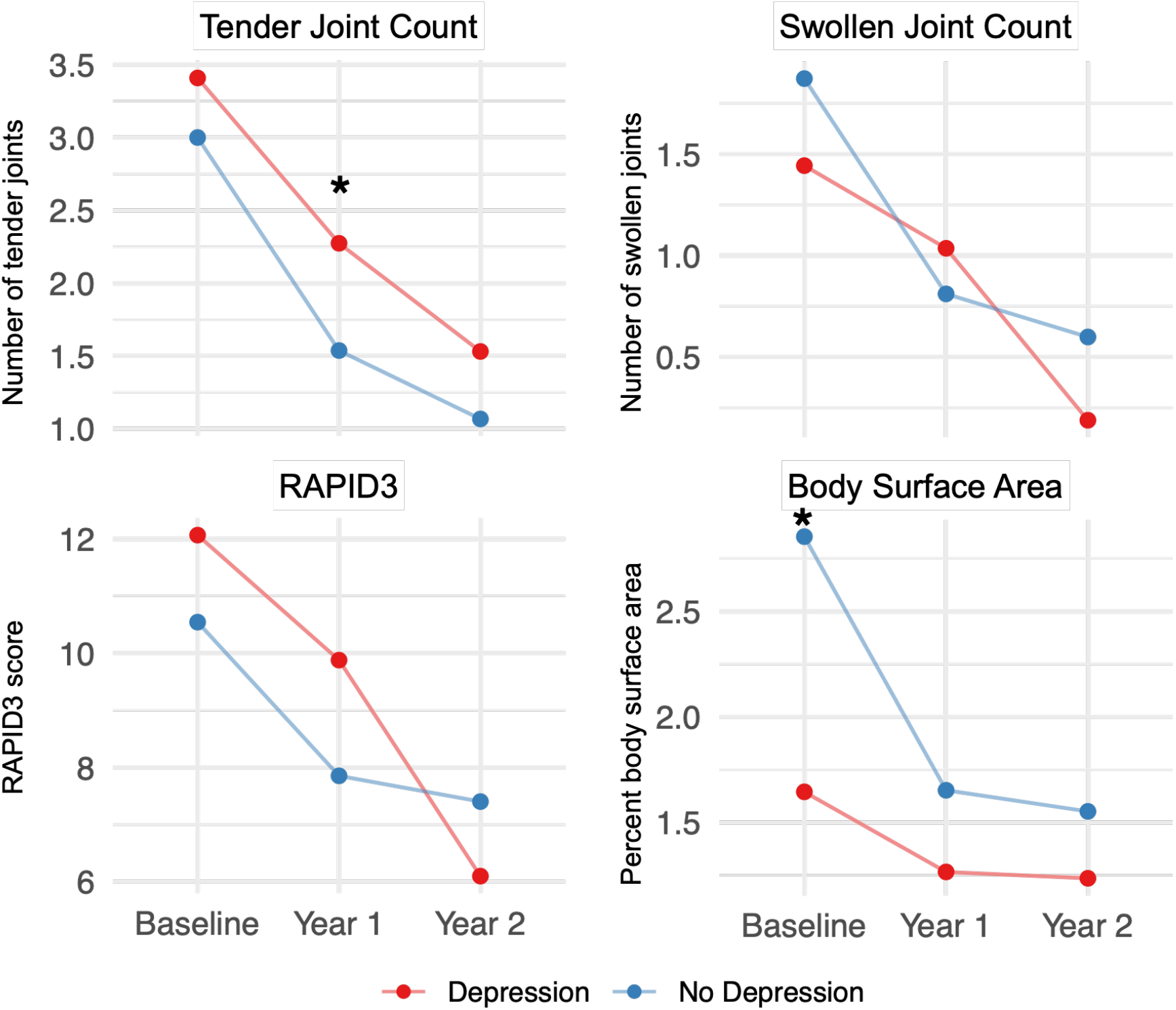
Observed mean PsA outcomes at baseline, year 1, and year 2. Red represents patients with depression, blue represents patients without depression. * p<0.05

When adjusting for age, sex, race, medication use, and comorbidities in the mixed effects regression models, patients with depression had numerically higher rate of TJC compared to those without depression (RR 1.23, 95%CI 0.78, 1.94, p=0.79) at baseline (Figure 2). This ratio was even higher at year 1 (RR 1.47, 95%CI 0.91, 2.35, p=0.19) and year 2 (RR 1.75, 95%CI 0.97, 3.14, p=0.07), nearing significance. This same pattern was not seen in the SJCs or in the difference estimates for RAPID3 and BSA.

**Figure 2.**
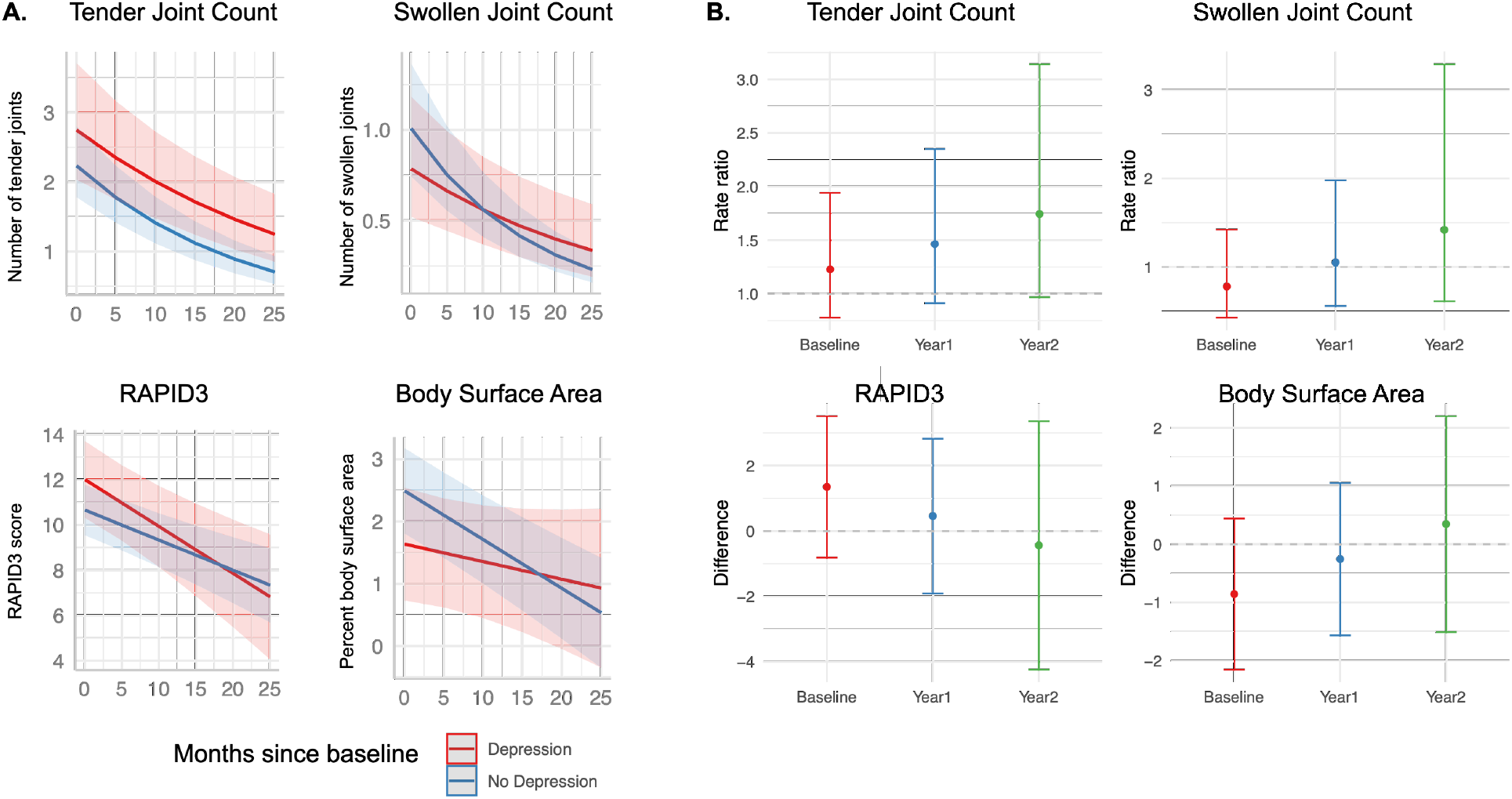
Model predicted means with 95% confidence bands (A), and predicted ratio ratios and estimated differences (b), adjusting for age, sex, race, comorbidities, and medication use.

## Discussion

We report on a longitudinal cohort of individuals with PsA seen at a tertiary care, combined-clinic setting in New York City. Observational cohort studies are of particular importance as they can inform on “real world” disease phenotypic characteristics and progression, which differ significantly from populations studied in the setting of controlled clinical trials. As in other recent reports, our cohort is composed by patients with a more oligoarticular, low skin disease involvement presentation. We also note a unique group of patients given its large referral base, urban setting, and diversity. While the NYU PAC population is primarily Caucasian, it has more racial and ethnic variability than any other previously described psoriasis and PsA cohorts^24^.

Importantly, this study emphasizes the high rates of depression and anxiety within the PsA population (22.8% and 18%, respectively). These numbers are in line with those reported by prior studies^15,25^. Of note, our study is the first to report rates of ADHD in an adult PsA population with a prevalence of 4%. This compares to world-wide rates of adults ranging from 1.1% to 4.4%^26-28^. Additionally, those who were depressed were more likely to be female and have cardiovascular disease, both of which are also seen in the general population^29,30^.

Despite the high prevalence of depression^31^ and anxiety^32^ and the fact that they have been independently associated with worse quality of life in psoriatic disease, these psychiatric disorders are often overlooked both in outcomes research and in clinical care. Here, we asked whether depression can affect PsA outcome measures. In so doing, we found that even in a population of relatively well controlled patients, over the course of treatment, while all patients improve, those with depression are less likely to see the same magnitude of amelioration in TJC compared to non-depressed patients, despite similar improvements in SJC. This implies that patient reported measures may be more affected by depression than physician observed measures.

Previous studies support our findings. Michelsen at al, looked at 728 patients with PsA and found that those with baseline depression and anxiety had increased patient global assessment of disease activity and joint pain^16^. These differences were largely driven by subjective outcomes, since depression and anxiety were not associated with inflammatory markers and swollen joint count during follow up. Freire et al showed that despite objective improvement on physician-assessed outcomes, patients with depression had increased pain VAS scores despite similar DAS28 and BASDAI scores, indicative of residual pain in these patients despite little objective disease activity and treatment^25^. Additionally, patients with RA and depression also had lower remission rates, driven by differences in pain score and patient global scores^33^.

In rheumatic diseases, depression and catastrophizing (i.e., the rumination and magnification of pain intensity) are associated with increased pain sensitivity, pain severity, and self-reported physical limitations^34-37^. This suggests that depression leads to a worsened subjective disease experience and, therefore, treatment outcomes. Our study supports this hypothesis and extends it to patients with PsA and depression. A possible mechanism behind this phenomenon may be related to how depression can lead to alterations in the central nervous system processing of pain such as through sensitization or the possible amplification of pain-related signals^36^. In individuals with RA, recent studies have shown that increased connectivity between the default mode network and the insula cortex was associated with increased disease activity, tender joints, and higher rates of centralized pain^38^. Additionally, in those with ankylosing spondylitis, functional brain MRI (fMRI) studies have shown that a higher degree of abnormal connectivity between the DMN and the salience network is related to higher pain intensity scores^39^. We propose that in a similar manner, neuroconnectivity in PsA patients is altered by compounded inflammation and psychological stress, triggering the pathways of sensory input and pain perception.

This is of particular importance given that many patients with PsA continue to have residual symptoms despite anti-inflammatory treatment. Even in patients with very low disease activity as defined by physician measures, up to half of them continue to report pain and fatigue^17^. This residual pain, despite seeming control of inflammation, constitutes a significant barrier to achieving remission while the presence of psychologic stress (i.e., depression) may be playing a contributing role in the persistence of this non-inflammatory type pain. Therefore, the current therapeutic paradigm of adjusting or escalating immunomodulatory medication may not be sufficient to mitigate pain in its entirety. Rather, patients may require supplementary approaches in treating their depression such as a more effective combination of antidepressant medications, introduction of cognitive behavioral therapy, or mindfulness interventions as co-adjuvant approaches to their standard of care PsA treatments.

Our study has several limitations, including the relatively short time course of data collection (i.e., four years) and the definition of “baseline” visit, which includes patients at different points during their disease journey. Patients are counted as having a diagnosis of depression or anxiety if they presented with an established diagnosis (patient report and/or ICD code) and/or use of psychiatric medication. This study did not capture the current presence or severity at the time the baseline visits or follow-ups.

In summary, despite similar improvements in more objective, physician-assessed outcomes (such as swollen joint count, enthesitis count, and percent of body surface area affected) compared to patients without depression, those with depression were less likely to experience the same amelioration in tender joint count and, to a lesser extent, RAPID3. This discrepancy is likely a manifestation of how depression may affect the way patients experience their PsA and their perception even after systemic therapies. Furthermore, it may explain why despite seemingly adequate inflammatory control and normalized physical exams, some patients continue to complain of pain.

Addressing underlying psychiatric comorbidities may bridge this gap between physician assessed measures and patient reported outcomes, and even reduce the need for escalation in therapy as residual pain may actually be inflammation-indepenedent. Therefore, depression should be considered a critical comorbidity when addressing PsA care in both clinical visits and in clinical trial settings. Our future research aims to address whether severity of depression symptoms and/or anti-depressant use can modulate PsA outcomes. This, coupled with investigations on neuroconnectivity dysregulation (as assessed by fMRIs) in PsA patients with comorbid depression as well as the incorporation of pharmacologic or non-pharmacologic modulation of psychiatric comorbidities may lead to improved understanding of residual pain and overall clinical outcomes in PsA.

## Data Availability

All data produced in the present study are available upon reasonable request to the authors

